# A Randomized, Double-Blinded, Placebo-Controlled, Phase 2 Study of Safety, Tolerability and Efficacy of Pirfenidone in Patients with Rheumatoid Arthritis Interstitial Lung Disease

**DOI:** 10.1101/2022.04.01.22273270

**Authors:** Joshua J. Solomon, Sonye Danoff, Felix Woodhead, Shelley Hurwitz, Rie Maurer, Ian Glaspole, Paul F. Dellaripa, Bibek Gooptu, Robert Vassallo, P. Gerald Cox, Kevin R. Flaherty, Huzaifa I. Adamali, Michael A. Gibbons, Lauren Troy, Ian Forrest, Joseph A. Lasky, Lisa G. Spencer, Jeffrey Golden, Mary Beth Scholand, Nazia Chaudhuri, Mark A. Perella, David Lynch, Daniel C. Chambers, Martin Kolb, Cathie Spino, Ganesh Raghu, Hilary Goldberg, Ivan O. Rosas, the TRAIL1 Network Investigators

## Abstract

**Background:** Interstitial lung disease (ILD) is a known complication of rheumatoid arthritis (RA) with a lifetime risk in any individual of 7.7%. The TRAIL1 trial was a randomized, double-blinded, placebo-controlled, phase 2 study of safety, tolerability, and efficacy of pirfenidone for the treatment of patients with RA-ILD.

**Methods:** The TRAIL1 was a phase 2 trial intended to enroll 270 adult patients (18 to 85 years) with established RA-ILD at 33 sites in 4 countries. Patients were randomly assigned (1:1) to 2,403 mg oral pirfenidone or placebo daily. The primary endpoint was the incidence of the composite endpoint of decline from baseline in percent predicted forced vital capacity (FVC%) of 10% or greater or death during the 52-week treatment period. Key secondary endpoints included change in absolute and FVC% over 52 weeks.

**Findings:** The trial was stopped early due to slow recruitment and soon after the shutdown of clinical trials as a consequence of the coronavirus disease 2019 (COVID-19) pandemic. Data from 123 patients enrolled were analyzed. The primary endpoint was met by 11.1% on pirfenidone vs. 15% on placebo [OR=0.67 (0.22, 2.03), p=0.48]. Subjects receiving pirfenidone had a slower rate of decline in lung function as measured by estimated annual change in FVC(ml) (−66 vs. -146, p=0.0082) and FVC(%) (−1.02 vs. -3.21, p=0.0028). This effect on decline was also seen when analyzed within participants with baseline usual interstitial pneumonia (UIP) pattern on HRCT (FVC(ml) (−43 vs. -169, p=0.0014) and FVC% (−0.2 vs. -3.81, p=0.0002)). There was no significant difference in the rate of treatment-emergent serious adverse events.

**Interpretation:** Due to early termination of the study, results should be interpreted with caution. Despite being underpowered to evaluate the primary endpoint, pirfenidone slowed the rate of decline of FVC over time in subjects with RA-ILD. Safety in patients with RA-ILD was similar to that seen in other pirfenidone trials.

**Funding:** Funding for this investigator initiated trial was provided by Genentech, Inc. to Ivan O. Rosas, MD, on behalf of the TRAIL1 Investigators.

## INTRODUCTION

Rheumatoid arthritis (RA) is the most common of the connective tissue diseases (CTD), affecting up to 0.75% of the United States (US) population, with increasing prevalence (1). The global prevalence of RA is 0.24% (or 16 million people), ranking as the 42^nd^ highest contributor to global disability (2). In the US alone, the annual excess health cost related to RA is estimated at $19.3 billion (3).

(4). While any lung compartment may be involved (5), interstitial lung disease (ILD) is the leading cause of excess morbidity and mortality amongst pulmonary complications in RA. The prevalence of RA-related ILD (RA-ILD) ranges from 19 to 63% (6), with a lifetime risk in any one patient of 7.7% (4). Usual interstitial pneumonia (UIP) pattern on Computed Tomography (CT) and pathology is the most common manifestation of RA-ILD and is associated with poor prognosis similar to patients with idiopathic pulmonary fibrosis (IPF). This distinguishes RA-ILD from many other forms of CTD-ILD, where non-specific interstitial pneumonia (NSIP) and organizing pneumonia (OP) are typically found. This similarity between RA-ILD and IPF has led to speculation that therapeutic agents with efficacy in IPF might also be beneficial in RA-ILD. Although scientific advances in the last decade have led to significant improvements in the control of the joint disease in RA, there is no known treatment for RA-ILD and thus the need for an efficacious treatment for RA-ILD is unmet to date.

Respiratory system involvement is common in RA and leads to increased morbidity and mortality

Pirfenidone is a therapeutic compound with both anti-inflammatory and anti-fibrotic properties. It is effective in several animal models of fibrosis (7) and has been studied in 15 controlled or uncontrolled clinical trials in human subjects with pulmonary fibrosis (8-22). It has been approved in many countries for the treatment of patients with IPF based on phase-3 clinical trial results (14). In the Treatment for Rheumatoid Arthritis and Interstitial Lung Disease 1 (TRAIL1) trial, we examined the effect of pirfenidone on the progression of lung disease in patients with RA and fibrotic ILD.

## METHODS/DESIGN

### Study Design and Oversight

The TRAIL1 study was a multinational randomized, double-blind, placebo-controlled, phase 2 study of the safety, tolerability, and efficacy of pirfenidone in patients with RA-ILD conducted in 33 sites in 4 countries (Australia, Canada, United Kingdom, and the US) (23). The trial was conducted in accordance with the trial protocol (available with the full text of this article) and the Harmonized Tripartite Guideline for Good Clinical Practice from the International Conference on Harmonization, and in compliance with the ethical principles of the Declaration of Helsinki. All participants provided written consent before entry into the study. Safety and regulatory oversight was conducted under the direction of a Data and Safety and Monitoring Board (DSMB), which was composed of individuals with expertise in RA-ILD, the study drugs and clinical trials research. At the completion of the study, authors had full access to the data for analysis.

### Patients

Recruitment began May 2017 and ended March 2020. Eligible patients were aged 18 to 85 years who were diagnosed with RA based on the 2010 American College of Rheumatology/European League Against Rheumatism (ACR/EULAR) criteria (24) and ILD based on high-resolution computed tomography (HRCT) and, when available, surgical lung biopsy. Screening HRCT scans required the presence of fibrotic abnormality affecting more than 10% of the lung parenchyma, with or without traction bronchiectasis or honeycombing and with no evidence or suspicion of an alternate diagnosis, as confirmed by a centrally adjudicated expert read. Patients were required to have a percent predicted forced vital capacity (FVC%) ≥ 40 and percent predicted diffusing capacity of the lung for carbon monoxide (DLCO%) ≥ 30 at screening as well as < 10% relative change in pre-bronchodilator FVC between the screening (Visit 1) and baseline study visits (Visit 2). Exclusion from participation included patients who had the introduction or dose alteration of corticosteroids or any cytotoxic, immuno-suppressive, cytokine-modulating or receptor antagonist agent specifically for the management of pulmonary manifestations of RA within 3 months of screening were excluded. In addition, patients with other lung manifestations of RA and those with a secondary CTD or overlap syndrome were excluded. All prohibited therapies (e.g., potent inhibitors or inducers of CYP1A2, etc.) must have been discontinued for at least 28 days before the start of screening (a comprehensive list of inclusion and exclusion criteria is provided in the Supplementary Appendix D).

### Study Design and Assessments

After written consent was obtained, eligible patients were randomized in a 1:1 ratio to receive oral pirfenidone (at a dose of 2,403 mg per day in divided doses) or placebo for 52 weeks (**Figure 1**). The study drug was titrated to full dose over a 14-day period and patients were maintained on study treatment for the duration of the trial (52 weeks). There were a total of 11 in-person visits, and the primary efficacy evaluation was conducted at week 52. Patients underwent spirometry and completed health-related quality of life (HrQOL) questionnaires at weeks 0, 13, 26, 39 and 52. HRCT scans were obtained at the beginning and end of the study period. All HRCT scans underwent a centralized interpretation to determine eligibility as well as pattern of ILD, and spirometry was centrally reviewed for adequacy and repeatability according to the American Thoracic Society criteria (25). The study protocol was approved by the institutional review board or ethics committee at each participating center and a DSMB maintained oversight throughout the trial. Details are provided in the Supplementary Appendix C.

**Figure 1.**
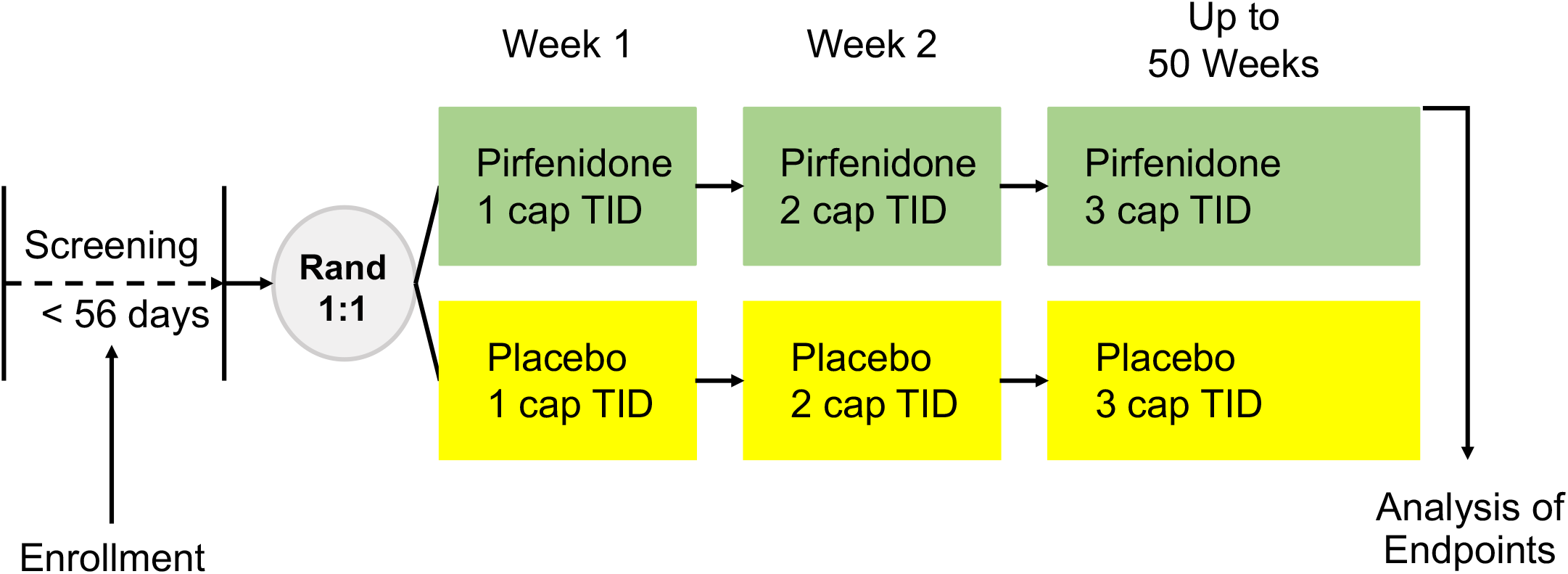
Trial Design.

### Endpoints

The primary endpoint of the study was the incidence of the composite endpoint of decline in percent predicted FVC of 10% or greater, or death, during the 52-week study period. Key secondary endpoints included changes in absolute and percent predicted FVC, the frequency of progressive disease (defined by the OMERACT (Outcome Measures in Rheumatoid Arthritis Clinical Trials) initiative as a categorical decline in FVC% and/or DLCO%) (26) and the change from baseline to week 52 in dyspnea, as measured by the Dyspnea-12 questionnaire (see Supplementary Appendix E for details).

Other pre-specified secondary endpoints include the rate of FVC change, time to the composite endpoint of 10% change in FVC or death, health outcomes (hospitalizations, mortality, adjudicated exacerbations, and transplant) and safety (adverse events (AEs) and serious adverse events (SAEs)). The incidence, type and severity of treatment-emergent adverse events were recorded and summarized according to primary System Organ Class and subcategorized by Medical Dictionary for Regulatory Activities preferred terms (version 19.1). Exploratory endpoints include measures of disease activity in RA (the Disease Activity Score (DAS), the Routine Assessment of Patient Index Data 3 score (RAPID3) and the Erythrocyte Sedimentation Rate (ESR)), biomarker expression, quantitative HRCT scores and Patient-Reported Outcomes (PROs) including the Leicester Cough Questionnaire (LCQ), the Patient Global Assessment and the Health Assessment Questionnaire.

### Statistical Analysis

The planned sample size was 270 participants randomized with equal probability to pirfenidone or placebo, to determine the outcomes of 254 participants with complete study data, providing at least 85% power to demonstrate the treatment arm difference for the primary endpoint. Enrollment was interrupted by the COVID-19 pandemic in March 2020. After centers were closed for approximately three months, and review of study feasibility with the DSMB, enrollment ended, and the study participants were followed per protocol. The final sample size was 123 participants with the last participant visit on April 7, 2021.

The primary efficacy outcome was the composite endpoint of a decline from baseline in FVC% of 10% or greater or death, analyzed using intention to treat principles. The screening spirometry values that met established quality criteria served as baseline randomization values and were performed before and after bronchodilator administration. The treatment arms were compared using a logistic regression model. Treatment arm was the main independent fixed effect, and pre-specified covariates included baseline percent predicted FVC and HRCT predominant pattern (UIP vs. other pattern). The alpha level for the primary endpoint was 0.05, two-sided, according to the trial design with no interim analyses. The adjusted odds ratio with 95% confidence interval was used to quantify the treatment effect for pirfenidone relative to placebo, with an odds ratio less than 1.0 indicating that pirfenidone is protective. A sensitivity analysis to assess the robustness of the primary analysis was performed using an adjusted per-protocol population with an expanded definition of treatment compliance.

Binary secondary endpoints, including annual rate of FVC decline in abslolute values (ml and % predicted) were analyzed using a logistic regression model similar to the primary endpoint analysis. Continuous, longitudinal secondary endpoints were analyzed using a restricted maximum likelihood-based repeated measures approach with random intercept. The longitudinal model included the effects of treatment arm, visit (actual weeks since randomization) and treatment arm-by-visit interaction, with the baseline value as covariate. The Kenward-Roger approximation was used and the covariance structure converging to the best fit by Akaike’s information criterion was selected. The analysis included all measurements obtained over the study period. The primary treatment comparison of slopes was assessed through the treatment-by-visit interaction.

Time-to-event secondary outcomes were analyzed using mixed effect proportional hazards regression modelling with treatment arm as main effect, network (US, UK, Canada, Australia) as random effect, and with baseline HRCT pattern (UIP vs. other patterns) as covariate. For time-to-event analyses involving spirometry, the baseline value was also a covariate. The censoring date was the earlier of the end of study period date or the date the participant was last observed without the event. Breslow’s method for handling ties was used, and Kaplan Meier plots and Schoenfeld residuals showed that the proportion hazards assumption was reasonable. The adjusted hazard ratio with 95% confidence interval was used to quantify the treatment effect for pirfenidone relative to placebo. For endpoints that indicate a decline in function, a hazard ratio less than 1.0 indicates that pirfenidone is protective.

Subgroup analyses related to the HRCT predominant pattern were not pre-specified. These were performed after observing a randomization imbalance with respect to the HRCT pattern, to generate hypotheses, and to allow comparison with relevant previously published trials that reported results for participants with UIP (27). Results related to the HRCT pattern subgroups were derived from full models described above, with the addition of a treatment by visit by HRCT pattern (UIP vs. other pattern) interaction term.

Exploratory endpoints, patient reported outcomes, adverse events, and variables related to administrative data were analyzed using chi square test for binary data or Wilcoxon-Mann-Whitney test for continuous data. The as-treated population was used for adverse events.

## RESULTS

### Study Patients

From May 2017 to March 2020, a total of 123 patients (63 in the pirfenidone group and 60 in the placebo group) were randomized (**Figure 2**). The study was terminated early due to slow enrollment and the sudden shutdown of routine clinical and research operational activities as a consequence of the global coronavirus disease 2019 (COVID-19) pandemic. Despite this abrupt cessation, all enrolled subjects completed the trial and there was no imputed data. With the exception of a higher percentage of subjects with a UIP pattern on HRCT in the placebo group, the demographics and baseline characteristics were similar and are summarized in **Table 1**. The majority of patients were male (60.3% and 60.5% in the pirfenidone and placebo groups, respectively), in their seventh decade of life (mean age of 66.6 and 68.1, respectively) and white (88.9% and 93.3%, respectively). The mean baseline percent predicted FVC and DLCO were 69.4% and 50% in the pirfenidone group and 70.4% and 47.6% in the placebo group. The mean CT extent of fibrosis was 20.8% and 24.2% respectively and the majority of patients had a UIP pattern on HRCT (54% and 78.3% in the pirfenidone and placebo groups, respectively). Among the patients who received at least one dose of study medication, 82.5% in the pirfenidone group and 85% in the placebo group completed the study. The duration of exposure to the study drug in the pirfenidone and placebo groups were similar (median (IQR) of 51.1 (19.1, 52.3) weeks and 51.4 (38.5, 52.2) weeks respectively) and the most frequent reasons for premature discontinuation of study drug were death and loss to follow-up (a total of 5 patients each in the intent-to-treat population).

**Table 1.** Demographics and Baseline Characteristics.

| Demographics | Pirfenidone<br>(N=63) | Placebo<br>(N=60) |
| --- | --- | --- |
| Age (years) | 66.6 ± 8.2 | 68.1 ± 9.1 |
| Sex |  |  |
| Female | 25 (39.7%) | 21 (35.0%) |
| Male | 38 (60.3%) | 39 (65.0%) |
| Race (n, %) |  |  |
| White | 56 (88.9%) | 56 (93.3%) |
| Black or African American | 2 (3.2%) | 1 (1.7%) |
| Asian | 4 (6.3%) | 0 (0.0%) |
| American Indian or Alaska Native | 1 (1.6%) | 0 (0.0%) |
| Native Hawaiian or Other Pacific<br>Islander | 0 (0.0%) | 0 (0.0%) |
| Multi-Race [1] | 0 (0.0%) | 0 (0.0%) |
| Unknown or Not Reported | 0 (0.0%) | 3 (5.0%) |
| Pulmonary Physiology |  |  |
| Percent Predicted FVC | 69.4 ± 14.8 | 70.4 ± 14.2 |
| FVC (L) | 2.6 ± 0.8 | 2.6 ± 0.8 |
| Percent Predicted DLCO | 50.0 ± 12.6 | 47.6 ± 12.8 |
| DLCO (mL/min/mmHg) | 12.0 ± 4.3 | 10.9 ± 4.4 |
| HRCT |  |  |
| CT extent of fibrosis, % (mean, SD) | 20.8 ± 9.8 | 24.2 ± 11.8 |
| Predominant HRCT pattern |  |  |
| UIP | 34 (54.0%) | 47 (78.3%) |
| NSIP | 9 (14.3%) | 4 (6.7%) |
| LIP | 0 (0.0%) | 3 (5.0%) |
| Indeterminate | 20 (31.7%) | 6 (10.0%) |
| Abbreviations: FVC=forced vital capacity, DLCO=diffusing capacity for carbon monoxide, HRCT=high-resolution computed tomography, SD=standard deviation, UIP=usual interstitial pneumonia, NSIP=non-specific interstitial pneumonia, LIP=lymphocytic interstitial pneumonia<br>Data are N (%) or mean and SD unless otherwise specified. Percentages are 100*n/N |  |  |

**Figure 2.**
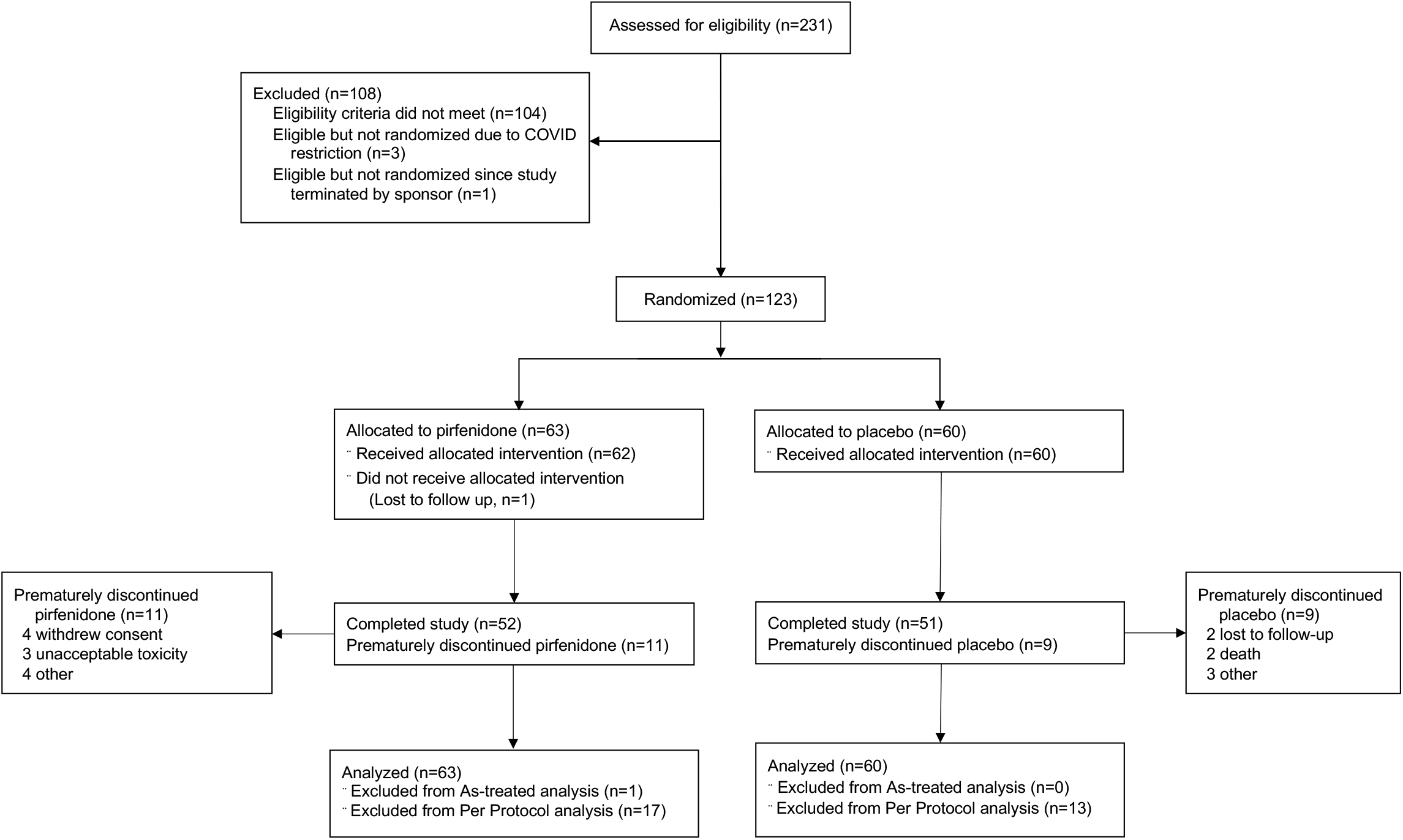
Trial Profile.

### Primary and Main Secondary Endpoints

There was no significant difference in patients who reached the primary, composite endpoint of a decline from baseline in percent predicted FVC of 10% or greater or death, analyzed using intention to treat principles (11.1% on pirfenidone vs. 15.0% on placebo; odds ratio [OR] 0.67 [95% CI 0.22 to 2.03], p=0.48) (**Table 2, Figure 3**). In pre-specified analysis of the change in FVC over 52 weeks, patients on pirfenidone had a slower rate of decline in lung function as measured by estimated annual change in FVC (ml) (−66 vs. -146, p=0.0082) (**Figure 4**). When patients were analyzed by HRCT pattern, the effect of pirfenidone on decline in FVC was more pronounced in subjects with UIP (−43 vs. -169, p=0.0014) (**Figure 5**).

**Table 2.** Primary and Secondary Efficacy Endpoints.

| Endpoint | Pirfenidone<br>(N=63) | Placebo<br>(N=60) | P value |
| --- | --- | --- | --- |
| **Primary Endpoint** |  |  |  |
| Decline in percent predicted FVC of<br>10% or death | 7 (11.1%) | 9 (15.0%) | 0.4823 |
| **Key Secondary Endpoints** |  |  |  |
| Decline in FVC - ml |  |  |  |
| Overall Population | -66 ± 21 | -146 ± 21 | 0.0082 |
| Patients with a UIP-pattern | -43 ± 31 | -169 ± 24 | 0.0014 |
| Decline in FVC - % |  |  |  |
| Overall Population | -1.02 ± 0.51 | -3.21 ± 0.52 | 0.0028 |
| Patients with a UIP-pattern | -0.20 ± 0.74 | -3.81 ± 0.70 | 0.0002 |
| **Other Secondary Endpoints** |  |  |  |
| Decline in percent predicted FVC by<br>10% or greater | 5 (7.9%) | 7 (11.7%) | 0.3235 |
| Frequency of progressive disease as<br>defined by OMERACT | 16 (25.4%) | 19 (31.7%) | 0.3531 |
| Change in Dyspnea-12 questionnaire | 0.46 ± 0.71 | 1.38 ± 0.72 | 0.3649 |
| All-cause mortality | 2 (3.2%) | 3 (5%) | 0.9930 |
| All-cause hospitalization | 7 (11.3%) | 7 (11.7%) | 0.6873 |
|  | 2 (3.2%) | 5 (8.3%) | 0.3484 |
| Hospitalizations for a respiratory<br>cause<br>Respiratory exacerbations requiring<br>hospitalization | 1 (1.6%) | 2 (3.3%) | 0.6157 |
| Abbreviations: FVC=forced vital capacity, UIP=usual interstitial pneumonia, OMERACT= Outcome Measures in Rheumatoid Arthritis Clinical Trials<br>Data are N (%) or adjusted mean and SE based on liner mixed model adjusting for baseline HRCT pattern and baseline values. Two-way interactions and three-way interaction were included for analysis of patients with a UIP-pattern, percentages are 100*n/N |  |  |  |

**Figure 3.**
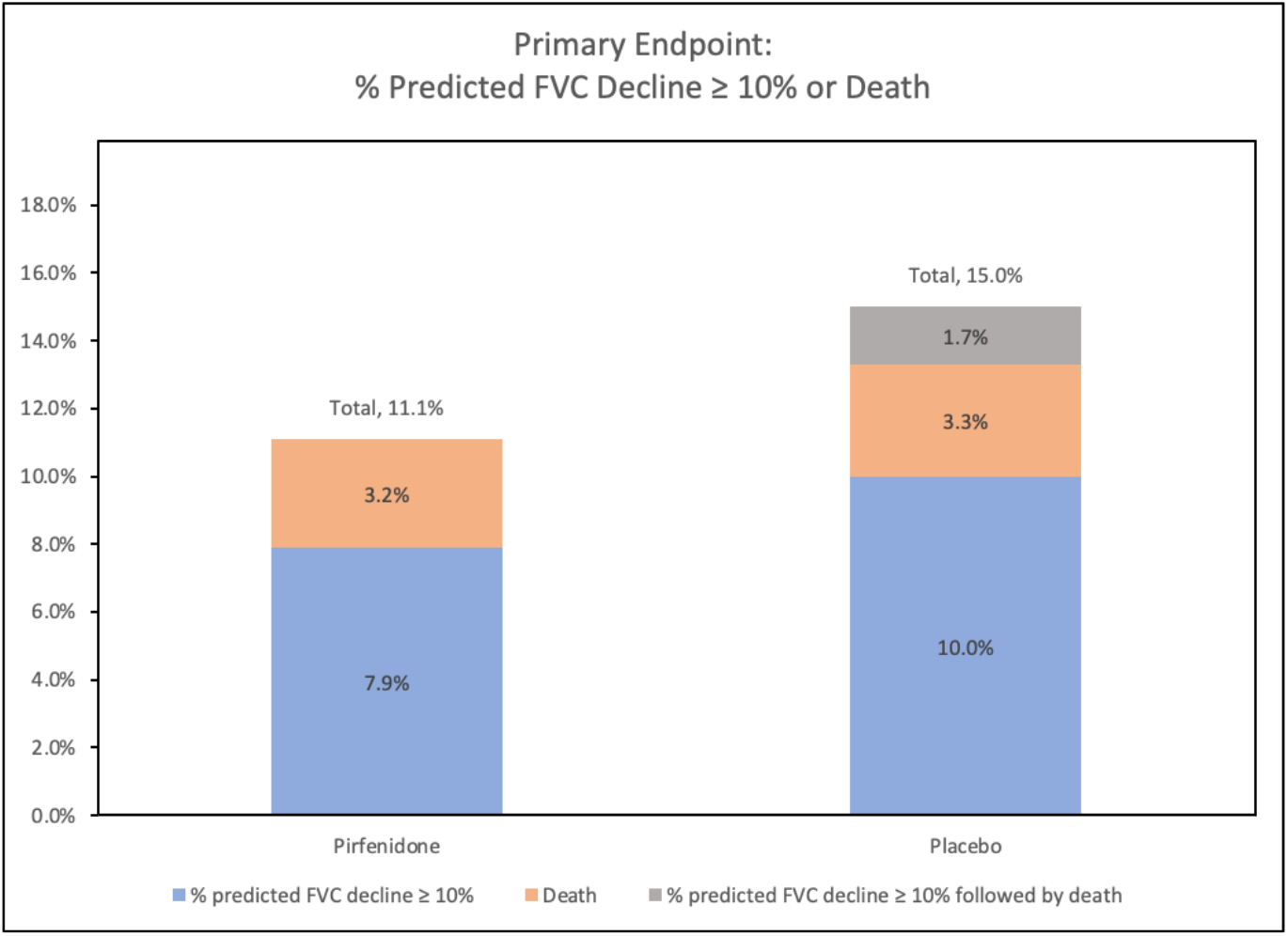
Decline from baseline in percent predicted FVC or greater or death over 52 weeks.

**Figure 4.**
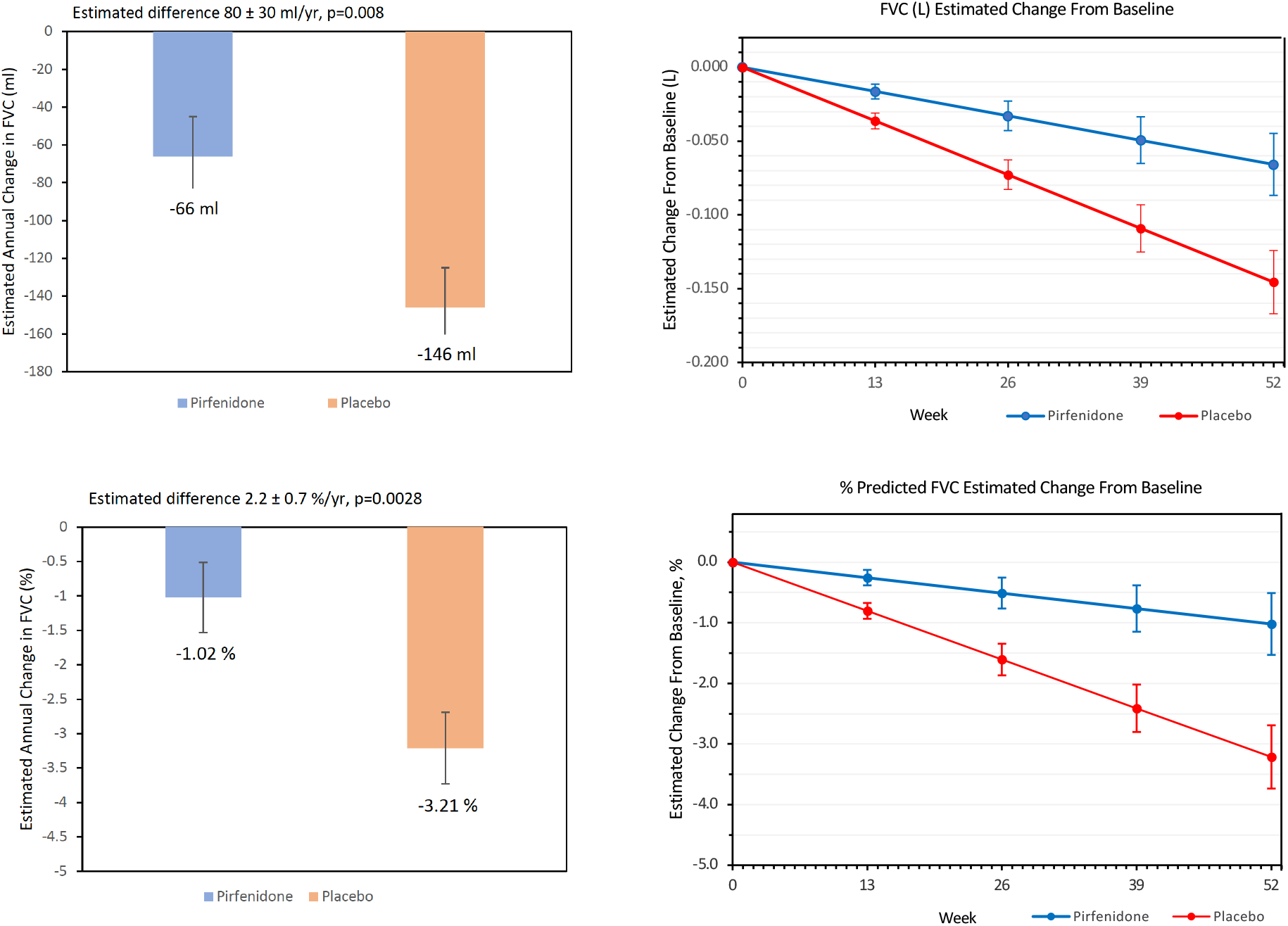
Estimated change in FVC (L, %) over 52 week by treatment group.

**Figure 5.**
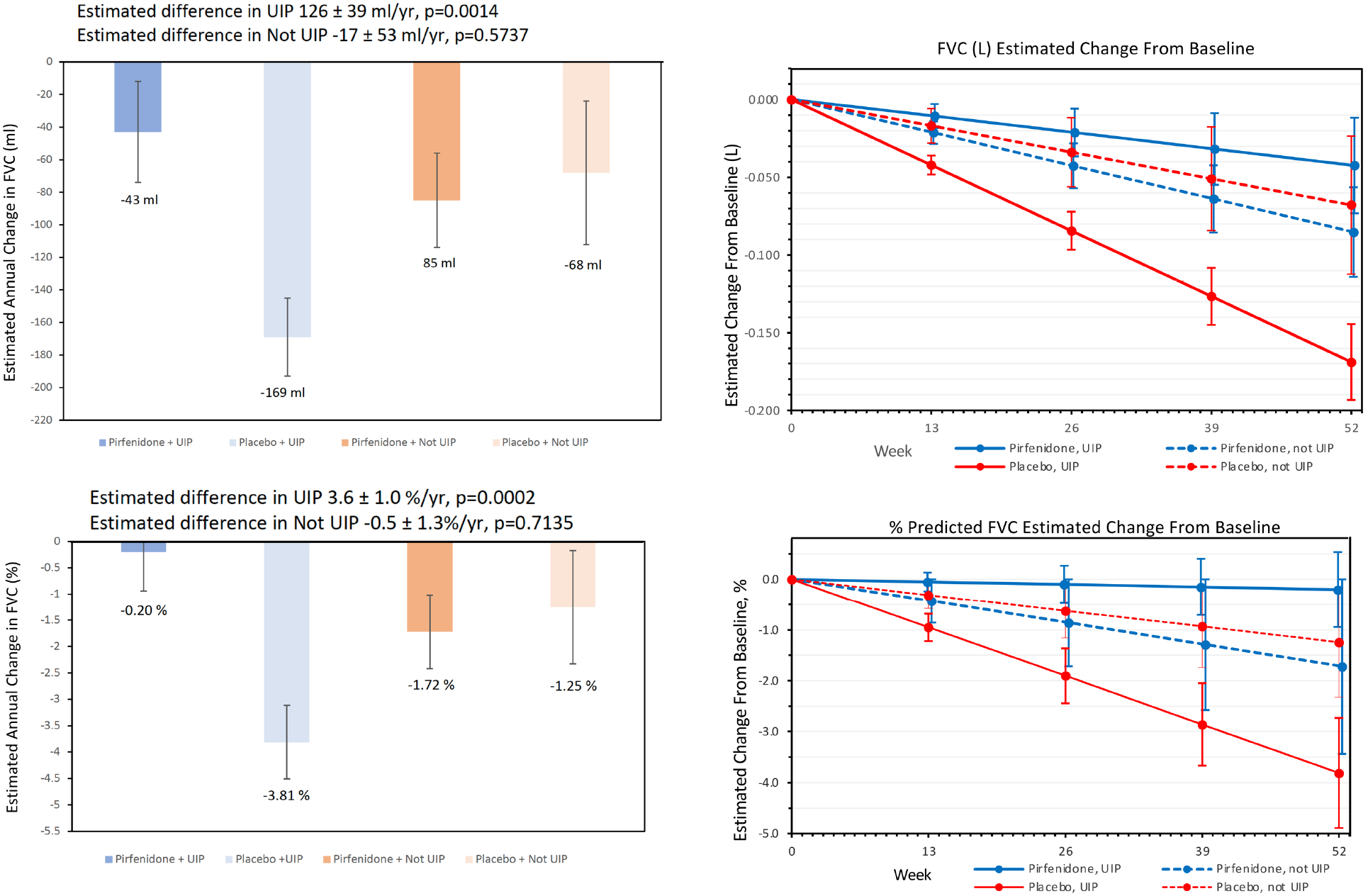
Estimated change in FVC (L, %) over 52 week by treatment group stratified by HRCT pattern.

### Other pre-specified Secondary Endpoints

In analysis of other secondary endpoints, the groups were similar with regards to the decline in percent predicted FVC by 10% or greater (7.9% vs. 11.7%, OR 0.52 (CI 0.143,1.898), p=0.32) and the frequency of progression as defined by OMERACT (25.4% vs. 31.7%, OR 0.678 (CI 0.299, 1.540), p=0.35). Hospitalizations and respiratory exacerbations were similar between the groups and there was no significant difference in all-cause mortality. There was no significant difference in change in Dyspnea-12 scores (0.46 in pirfenidone vs. 1.38 in placebo, p=0.36).

### Adverse Events

Adverse events are summarized in **Table 3**. Treatment-emergent adverse events were seen in 100% of subjects on pirfenidone and 93.3% of subjects on placebo (p=0.0387). Of these, adverse events felt to be related to treatment were reported more frequently in the pirfenidone group (43.6% vs 30.0%, p<0.1210). The most frequent adverse events, analyzed in the as-treated population, were nausea, fatigue, and diarrhea. These adverse events were generally mild and not clinically significant.

**Table 3.** Adverse Events

|  | Pirfenidone<br>(N=62) | Placebo<br>(N=60) | P value |
| --- | --- | --- | --- |
| Treatment-emergent adverse events | 62 (100%) | 56 (93.3%) | 0.0387 |
| Treatment-emergent serious adverse events | 9 (14.5%) | 8 (13.3%) | 0.8504 |
| Treatment-emergent/treatment-related adverse events | 27 (43.6%) | 18 (30.0%) | 0.1210 |
| Nausea | 33 (53.2%) | 11 (18.3%) | <0.0001 |
| Fatigue | 20 (32.3%) | 12 (20.0%) | 0.1239 |
| Diarrhea | 19 (30.7%) | 16 (26.7%) | 0.6272 |
| Cough | 18 (29.0%) | 12 (20.0%) | 0.2468 |
| Headache | 18 (29.0%) | 8 (13.3%) | 0.0343 |
| Anorexia | 17 (27.4%) | 6 (10.0%) | 0.0139 |
| Breathlessness | 13 (21.0%) | 11 (18.3%) | 0.7144 |
| GERD | 13 (21.0%) | 6 (10.0%) | 0.0949 |
| Vomiting | 12 (19.4%) | 4 (6.7%) | 0.0585 |
| Joint Pain | 11 (17.7%) | 13 (21.7%) | 0.5856 |
| Treatment-emergent/treatment-related serious adverse events | 1 (1.6%) | 0 (0.0%) | 0.3233 |
| Adverse events leading to discontinuation of study drug | 15 (24.2%) | 6 (10.0%) | 0.0379 |
| Treatment-emergent death or transplant | 2 (3.2%) | 4 (6.7%) | 0.4355 |
| Treatment-emergent RA-ILD-related mortality | 1 (1.6%) | 0 (0.0%) | 1.0 |
| Abbreviations: GERD=gastroesophageal reflux disease, RA-ILD=rheumatoid arthritis-associated interstitial lung disease<br>Data are N (%). Percentages are 100*n/N |  |  |  |

There was no difference between treatment arms in the number of treatment-related serious adverse events. Adverse events leading to the discontinuation of study drug occurred more frequently in those on pirfenidone (24% vs. 10%, p=0.0379). There was a total of 5 deaths (2 in the pirfenidone group and 3 in the placebo group), none of which were felt to be secondary to study drug. One of the 5 deaths was felt to be secondary to RA-ILD. There were no new safety signals identified. When analyses were limited to subjects on background therapy for RA (anti-inflammatories, TNF agents and rituximab), there was no difference between treatment arms in adverse events and serious adverse events (see Supplementary Appendix Table S2).

## DISCUSSION

In the TRAIL1 study we sought to investigate the safety and efficacy of pirfenidone in subjects with RA and fibrotic ILD. The trial was unfortunately aborted as the consequence of the global pandemic and anticipated failure to enroll the planned sample size and was therefore underpowered to evaluate the primary endpoint and draw meaningful conclusion. However, pre-specified secondary endpoints revealed a reduction in the decline in FVC over 52 weeks in those patients on pirfenidone relative to placebo. Though not statistically significant, differences in other secondary endpoints such as categorical decline in FVC%, frequency of progressive disease, change in the Dyspnea-12 and hospitalizations from a respiratory cause all favored pirfenidone. The magnitude of effect of pirfenidone on FVC decline in RA-ILD was similar to that seen in IPF and non-IPF progressive fibrotic ILDs in the ASCEND, INBUILD, pirfenidone in progressive fibrosing unclassifiable ILD and RELIEF trials (14, 28, 29). Importantly, there were no new safety signals identified and treatment was well tolerated.

Recruitment for the TRAIL1 trial was challenging. Acknowledging that we were behind in recruitment prior to the COVID-19 pandemic, the abrupt shutdown of routine clinical and research activities as a consequence of the pandemic, and prioritization of the need to attend to management of COVID-19 patients at all centers worldwide led to the decision with input from the DSMB to terminate further enrollment. Power calculations determined that a sample size of 270 participants were needed to demonstrate the treatment arm difference for the primary endpoint. Recruitment was terminated at 123 participants and all subjects finished the trial with no data imputation. Despite enrolling less than half of the originally planned cohort, pirfenidone reduced the FVC decline in all subjects by 55% and by 75% in those with a UIP-pattern of fibrosis.

Limited data suggest the influence of ILD on quality of life in those with RA is substantial, with patients scoring the same or worse on quality-of-life measures as those with IPF, a progressive fatal fibrosing ILD (30). RA-ILD is a progressive disease; 60% show radiographic progression over an 18-month period (31). ILD contributes to mortality in 7 to 9% of patients with RA (32) and patients with RA-ILD have an average lifespan of 2.6 years compared to 10 years in age-matched RA patients without ILD (4). UIP is the most common radiographic pattern of lung injury (33) which is associated with a worse outcome (34-36). In addition to its impact on patients, the burden of RA-ILD on the healthcare system is significant. The presence of ILD leads to an additional estimated 5-year cost of US $173,000 per patient (37).

RA-ILD holds many similarities to IPF, with shared risk factors of male sex, older age and history of smoking (6, 38). The HRCT appearance of UIP in RA and IPF is frequently indistinguishable. Other notable similarities include overlapping serum biomarker profiles (e.g., Krebs von den Lungen-6 (KL-6 (39, 40)), shared genetic risk factors (MUC5B and telomerase mutations (41-44)) and serologic evidence of autoimmunity (45). RA-UIP and IPF have previously been shown to confer a similar poor prognosis (35, 36) though more recent studies have suggested a better prognosis in RA-ILD (46). These similarities and the proven benefit of pirfenidone on IPF (14) and other fibrotic ILDs that progress despite therapy (28) led us to evaluate the safety and efficacy of pirfenidone in RA-ILD.

Our inclusion criteria selected for those with a fibrotic subtype of ILD in RA, with the mean extent of fibrosis as scored by a radiologist of ≥ 20% in both groups and restrictive physiology. CT extent of fibrotic disease has been associated with mortality in IPF (47-49) but has not been thoroughly evaluated in RA-ILD. Our placebo group had an average FVC decline of 146 ml over 52 weeks. Though markedly less than subjects with IPF in the placebo groups in ASCEND (428 ml/yr) (14) and INPULSIS (239.9 ml/yr and -207.3 ml/yr in INPULSIS 1 and 2 respectively) (50), this decline is closer to that seen in INBUILD (187.8 ml/yr in all subjects (27) and 199.3 ml/yr in subjects with RA-ILD (51)) and RELIEF (114.4 ml/yr) (52). These two trials enrolled subjects with evidence of progression prior to enrollment, and similar progression over time in our placebo group supports the idea that RA with fibrotic ILD is a progressive fibrosing interstitial lung disease.

The effect of treatment with pirfenidone on the decline in FVC was more pronounced in those with a UIP pattern on HRCT (for the overall population, 11.1% in the treated group vs. 15% in the overall population reached the primary endpoint, compared to 8.8% vs. 14.9% in those with a UIP pattern on HRCT). Of the 16 participants who died or were hospitalized, 14 had a UIP pattern on imaging. Though the post-hoc nature of this analysis limits generalizability, these findings suggest that the pattern of fibrosis could have therapeutic and prognostic significance above that provided by extent of fibrosis and deserves further study.

Treatment with pirfenidone was safe and well tolerated. Side effects were similar to those seen in other studies of pirfenidone in patients with ILD (14, 29). The most common side effect was nausea, seen in half of patients on pirfenidone. Though the number of serious adverse events were lower in our study compared to ASCEND (14.5% vs. 18.7% respectively), subjects with RA-ILD had a higher rate of study-drug discontinuation (24.2% in TRAIL1 vs. 14.4% in ASCEND). As opposed to IPF, patients with RA-ILD have multi-system disease and are often on disease-modifying antirheumatic drugs (DMARDs) with their own side effect profile. When adverse and severe adverse events were analyzed in those on background agents for RA, no differences were noted between pirfenidone and placebo. The ability of pirfenidone to be added to background DMARD therapy for patients with RA in a safe and tolerable fashion is a significant finding of our study.

In spite of the positive finding of a slower decline in FVC in patients treated with pirfenidone, and acknowledging the early termination of the trial, we surface the following limitations: 1) due to early termination, we failed to meet the primary endpoint and results should be interpreted with care, 2) selection bias with an inclusion criteria of least 10% fibrosis on HRCT and an unknown applicability of our findings to patients with less fibrosis on HRCT and finally, 3) there were more patients with UIP in the placebo arm of our trial though UIP pattern was used as a pre-established covariate in our analyses.

In summary, we present results from the first randomized placebo-controlled trial conducted exclusively in subjects with RA-ILD. We were forced to terminate the trial due to unforeseen circumstances that included the abrupt consequences of the COVID-19 pandemic and were underpowered to meet our primary endpoint. However, the use of pirfenidone was associated with a slower rate of FVC% decline in patients with RA-ILD compared to placebo and the totality of the evidence suggests that pirfenidone is effective in the treatment of RA-ILD. Treatment was well tolerated despite background RA-related therapy and no new safety signals were identified. In addition, we identified a subgroup of RA-ILD patients at risk for significant progression over time. UIP is the most common radiographic pattern seen in patients with RA-ILD and our data suggest they have a faster progression and may have a more robust response to therapy. Future studies with anti-fibrotics should stratify cohorts of patients with RA-ILD patients to those with a UIP pattern and non UIP, fibrotic ILD on HRCT.

## Supporting information

Complete supplement

## Data Availability

TRAIL1 network will share de-identified study information to approved investigators

https://www.clinicaltrials.gov

## Acknowledgments

The TRAIL1 study team (listed in the Supplement) thanks and acknowledges all patients and their referring physicians for their participation in this important research; as well as the members of the Data Safety and Monitoring Board for their participation.

## REFERENCES

1. Hunter TM, Boytsov NN, Zhang X, Schroeder K, Michaud K, Araujo AB. Prevalence of rheumatoid arthritis in the United States adult population in healthcare claims databases, 2004-2014. Rheumatol Int. 2017;37(9):1551–7. Epub 2017/04/30. doi: 10.1007/s00296-017-3726-1. PubMed PMID: 28455559.

2. Cross M, Smith E, Hoy D, Carmona L, Wolfe F, Vos T, et al. The global burden of rheumatoid arthritis: estimates from the global burden of disease 2010 study. Ann Rheum Dis. 2014;73(7):1316–22. doi: 10.1136/annrheumdis-2013-204627. PubMed PMID: 24550173.

3. Birnbaum H, Pike C, Kaufman R, Marynchenko M, Kidolezi Y, Cifaldi M. Societal cost of rheumatoid arthritis patients in the US. Curr Med Res Opin. 2010;26(1):77–90. doi: 10.1185/03007990903422307. PubMed PMID: 19908947.

4. Bongartz T, Nannini C, Medina-Velasquez YF, Achenbach SJ, Crowson CS, Ryu JH, et al. Incidence and mortality of interstitial lung disease in rheumatoid arthritis: a population-based study. Arthritis Rheum. 2010;62(6):1583–91. Epub 2010/02/16. doi: 10.1002/art.27405. PubMed PMID: 20155830.

5. Brown KK. Rheumatoid lung disease. Proc Am Thorac Soc. 2007;4(5):443–8. Epub 2007/08/09. doi: 4/5/443 [pii] 10.1513/pats.200703-045MS. PubMed PMID: 17684286; PubMed Central PMCID: PMC2647595.

6. Demoruelle MK, Solomon JJ, Olson AL. The Epidemiology of Rheumatoid Arthritis-Associated Lung Disease. In: Fischer A, Lee JS, editors. Lung Disease in Rheumatoid Arthritis. 1 ed: Humana Press; 2018. p. 45–58.

7. Schaefer CJ, Ruhrmund DW, Pan L, Seiwert SD, Kossen K. Antifibrotic activities of pirfenidone in animal models. Eur Respir Rev. 2011;20(120):85–97. Epub 2011/06/03. doi: 10.1183/09059180.00001111. PubMed PMID: 21632796.

8. Nagai S, Hamada K, Shigematsu M, Taniyama M, Yamauchi S, Izumi T. Open-label compassionate use one year-treatment with pirfenidone to patients with chronic pulmonary fibrosis. Intern Med. 2002;41(12):1118–23. Epub 2003/01/11. PubMed PMID: 12521199.

9. Raghu G, Johnson WC, Lockhart D, Mageto Y. Treatment of idiopathic pulmonary fibrosis with a new antifibrotic agent, pirfenidone: results of a prospective, open-label Phase II study. Am J Respir Crit Care Med. 1999;159(4 Pt 1):1061–9. Epub 1999/04/08. doi: 10.1164/ajrccm.159.4.9805017. PubMed PMID: 10194146.

10. Gahl WA, Brantly M, Troendle J, Avila NA, Padua A, Montalvo C, et al. Effect of pirfenidone on the pulmonary fibrosis of Hermansky-Pudlak syndrome. Mol Genet Metab. 2002;76(3):234–42. Epub 2002/07/20. PubMed PMID: 12126938.

11. Azuma A, Nukiwa T, Tsuboi E, Suga M, Abe S, Nakata K, et al. Double-blind, placebo-controlled trial of pirfenidone in patients with idiopathic pulmonary fibrosis. Am J Respir Crit Care Med. 2005;171(9):1040–7. Epub 2005/01/25. doi: 10.1164/rccm.200404-571OC. PubMed PMID: 15665326.

12. Taniguchi H, Ebina M, Kondoh Y, Ogura T, Azuma A, Suga M, et al. Pirfenidone in idiopathic pulmonary fibrosis. Eur Respir J. 2010;35(4):821–9. Epub 2009/12/10. doi: 10.1183/09031936.00005209. PubMed PMID: 19996196.

13. Noble PW, Albera C, Bradford WZ, Costabel U, Glassberg MK, Kardatzke D, et al. Pirfenidone in patients with idiopathic pulmonary fibrosis (CAPACITY): two randomised trials. Lancet. 2011;377(9779):1760–9. Epub 2011/05/17. doi: 10.1016/S0140-6736(11)60405-4. PubMed PMID: 21571362.

14. King TE, Jr., Bradford WZ, Castro-Bernardini S, Fagan EA, Glaspole I, Glassberg MK, et al. A phase 3 trial of pirfenidone in patients with idiopathic pulmonary fibrosis. N Engl J Med. 2014;370(22):2083–92. Epub 2014/05/20. doi: 10.1056/NEJMoa1402582. PubMed PMID: 24836312.

15. Huang H, Dai HP, Kang J, Chen BY, Sun TY, Xu ZJ. Double-Blind Randomized Trial of Pirfenidone in Chinese Idiopathic Pulmonary Fibrosis Patients. Medicine (Baltimore). 2015;94(42):e1600. Epub 2015/10/27. doi: 10.1097/MD.0000000000001600. PubMed PMID: 26496265; PubMed Central PMCID: PMCPMC4620844.

16. Costabel U, Albera C, Bradford WZ, Hormel P, King TE, Jr., Noble PW, et al. Analysis of lung function and survival in RECAP: An open-label extension study of pirfenidone in patients with idiopathic pulmonary fibrosis. Sarcoidosis Vasc Diffuse Lung Dis. 2014;31(3):198–205. Epub 2014/11/05. PubMed PMID: 25363219.

17. Ogura T, Taniguchi H, Azuma A, Inoue Y, Kondoh Y, Hasegawa Y, et al. Safety and pharmacokinetics of nintedanib and pirfenidone in idiopathic pulmonary fibrosis. Eur Respir J. 2015;45(5):1382–92. Epub 2014/12/17. doi: 10.1183/09031936.00198013. PubMed PMID: 25504994.

18. Behr J, Bendstrup E, Crestani B, Gunther A, Olschewski H, Skold CM, et al. Safety and tolerability of acetylcysteine and pirfenidone combination therapy in idiopathic pulmonary fibrosis: a randomised, double-blind, placebo-controlled, phase 2 trial. Lancet Respir Med. 2016;4(6):445–53. Epub 2016/05/11. doi: 10.1016/S2213-2600(16)30044-3. PubMed PMID: 27161257.

19. Khanna D, Albera C, Fischer A, Khalidi N, Raghu G, Chung L, et al. An Open-label, Phase II Study of the Safety and Tolerability of Pirfenidone in Patients with Scleroderma-associated Interstitial Lung Disease: the LOTUSS Trial. J Rheumatol. 2016;43(9):1672–9. Epub 2016/07/03. doi: 10.3899/jrheum.151322. PubMed PMID: 27370878.

20. Iwata T, Yoshino I, Yoshida S, Ikeda N, Tsuboi M, Asato Y, et al. A phase II trial evaluating the efficacy and safety of perioperative pirfenidone for prevention of acute exacerbation of idiopathic pulmonary fibrosis in lung cancer patients undergoing pulmonary resection: West Japan Oncology Group 6711 L (PEOPLE Study). Respir Res. 2016;17(1):90. Epub 2016/07/28. doi: 10.1186/s12931-016-0398-4. PubMed PMID: 27450274; PubMed Central PMCID: PMCPMC4957367.

21. Vancheri C, Kreuter M, Richeldi L, Ryerson CJ, Valeyre D, Grutters JC, et al. Nintedanib with Add-on Pirfenidone in Idiopathic Pulmonary Fibrosis. Results of the INJOURNEY Trial. Am J Respir Crit Care Med. 2018;197(3):356–63. Epub 2017/09/12. doi: 10.1164/rccm.201706-1301OC. PubMed PMID: 28889759.

22. Costabel U, Albera C, Lancaster LH, Lin CY, Hormel P, Hulter HN, et al. An Open-Label Study of the Long-Term Safety of Pirfenidone in Patients with Idiopathic Pulmonary Fibrosis (RECAP). Respiration. 2017;94(5):408–15. Epub 2017/09/13. doi: 10.1159/000479976. PubMed PMID: 28898890.

23. Solomon JJ, Danoff SK, Goldberg HJ, Woodhead F, Kolb M, Chambers DC, et al. The Design and Rationale of the Trail1 Trial: A Randomized Double-Blind Phase 2 Clinical Trial of Pirfenidone in Rheumatoid Arthritis-Associated Interstitial Lung Disease. Adv Ther. 2019;36(11):3279–87. Epub 2019/09/14. doi: 10.1007/s12325-019-01086-2. PubMed PMID: 31515704.

24. Aletaha D, Neogi T, Silman AJ, Funovits J, Felson DT, Bingham CO, 3rd, et al. 2010 Rheumatoid arthritis classification criteria: an American College of Rheumatology/European League Against Rheumatism collaborative initiative. Arthritis Rheum. 2010;62(9):2569–81. doi: 10.1002/art.27584. PubMed PMID: 20872595.

25. Miller MR, Hankinson J, Brusasco V, Burgos F, Casaburi R, Coates A, et al. Standardisation of spirometry. Eur Respir J. 2005;26(2):319–38. Epub 2005/08/02. doi: 10.1183/09031936.05.00034805. PubMed PMID: 16055882.

26. Khanna D, Mittoo S, Aggarwal R, Proudman SM, Dalbeth N, Matteson EL, et al. Connective Tissue Disease-associated Interstitial Lung Diseases (CTD-ILD) - Report from OMERACT CTD-ILD Working Group. J Rheumatol. 2015;42(11):2168–71. Epub 2015/03/03. doi: 10.3899/jrheum.141182. PubMed PMID: 25729034; PubMed Central PMCID: PMCPMC4809413.

27. Flaherty KR, Wells AU, Cottin V, Devaraj A, Walsh SLF, Inoue Y, et al. Nintedanib in Progressive Fibrosing Interstitial Lung Diseases. N Engl J Med. 2019;381(18):1718–27. Epub 2019/10/01. doi: 10.1056/NEJMoa1908681. PubMed PMID: 31566307.

28. Behr J, Neuser P, Prasse A, Kreuter M, Rabe K, Schade-Brittinger C, et al. Exploring efficacy and safety of oral Pirfenidone for progressive, non-IPF lung fibrosis (RELIEF) - a randomized, double-blind, placebo-controlled, parallel group, multi-center, phase II trial. BMC Pulm Med. 2017;17(1):122. Epub 2017/09/08. doi: 10.1186/s12890-017-0462-y. PubMed PMID: 28877715; PubMed Central PMCID: PMCPMC5588600.

29. Maher TM, Corte TJ, Fischer A, Kreuter M, Lederer DJ, Molina-Molina M, et al. Pirfenidone in patients with unclassifiable progressive fibrosing interstitial lung disease: a double-blind, randomised, placebo-controlled, phase 2 trial. Lancet Respir Med. 2020;8(2):147–57. Epub 2019/10/04. doi: 10.1016/S2213-2600(19)30341-8. PubMed PMID: 31578169.

30. Natalini JG, Swigris JJ, Morisset J, Elicker BM, Jones KD, Fischer A, et al. Understanding the determinants of health-related quality of life in rheumatoid arthritis-associated interstitial lung disease. Respir Med. 2017;127:1–6. doi: 10.1016/j.rmed.2017.04.002. PubMed PMID: 28502413.

31. Gochuico BR, Avila NA, Chow CK, Novero LJ, Wu HP, Ren P, et al. Progressive preclinical interstitial lung disease in rheumatoid arthritis. Arch Intern Med. 2008;168(2):159–66. Epub 2008/01/30. doi: 10.1001/archinternmed.2007.59. PubMed PMID: 18227362.

32. Olson AL, Swigris JJ, Sprunger DB, Fischer A, Fernandez-Perez ER, Solomon J, et al. Rheumatoid arthritis-interstitial lung disease-associated mortality. Am J Respir Crit Care Med. 2011;183(3):372–8. Epub 2010/09/21. doi: 10.1164/rccm.201004-0622OC. PubMed PMID: 20851924.

33. Tanaka N, Kim JS, Newell JD, Brown KK, Cool CD, Meehan R, et al. Rheumatoid arthritis-related lung diseases: CT findings. Radiology. 2004;232(1):81–91. Epub 2004/05/29. doi: 10.1148/radiol.2321030174. PubMed PMID: 15166329.

34. Kim EJ, Elicker BM, Maldonado F, Webb WR, Ryu JH, Van Uden JH, et al. Usual interstitial pneumonia in rheumatoid arthritis-associated interstitial lung disease. Eur Respir J. 2010;35(6):1322–8. Epub 2009/12/10. doi: 09031936.00092309 [pii] 10.1183/09031936.00092309. PubMed PMID: 19996193.

35. Park JH, Kim DS, Park IN, Jang SJ, Kitaichi M, Nicholson AG, et al. Prognosis of fibrotic interstitial pneumonia: idiopathic versus collagen vascular disease-related subtypes. Am J Respir Crit Care Med. 2007;175(7):705–11. Epub 2007/01/16. doi: 200607-912OC [pii] 10.1164/rccm.200607-912OC. PubMed PMID: 17218621.

36. Solomon JJ, Ryu JH, Tazelaar HD, Myers JL, Tuder R, Cool CD, et al. Fibrosing interstitial pneumonia predicts survival in patients with rheumatoid arthritis-associated interstitial lung disease (RA-ILD). Respir Med. 2013;107(8):1247–52. doi: 10.1016/j.rmed.2013.05.002. PubMed PMID: 23791462.

37. Raimundo K, Solomon JJ, Olson AL, Kong AM, Cole AL, Fischer A, et al. Rheumatoid Arthritis–Interstitial Lung Disease in the United States: Prevalence, Incidence, and Healthcare Costs and Mortality. The Journal of Rheumatology. 2018. doi: 10.3899/jrheum.171315.

38. American Thoracic Society. Idiopathic pulmonary fibrosis: diagnosis and treatment. International consensus statement. American Thoracic Society (ATS), and the European Respiratory Society (ERS). Am J Respir Crit Care Med. 2000;161(2 Pt 1):646–64. doi: 10.1164/ajrccm.161.2.ats3-00.

39. Lee JS, Lee EY, Ha YJ, Kang EH, Lee YJ, Song YW. Serum KL-6 levels reflect the severity of interstitial lung disease associated with connective tissue disease. Arthritis Res Ther. 2019;21(1):58. Epub 2019/02/16. doi: 10.1186/s13075-019-1835-9. PubMed PMID: 30764869; PubMed Central PMCID: PMCPMC6376648.

40. Yokoyama A, Kondo K, Nakajima M, Matsushima T, Takahashi T, Nishimura M, et al. Prognostic value of circulating KL-6 in idiopathic pulmonary fibrosis. Respirology. 2006;11(2):164–8. doi: 10.1111/j.1440-1843.2006.00834.x. PubMed PMID: 16548901.

41. Juge PA, Lee JS, Ebstein E, Furukawa H, Dobrinskikh E, Gazal S, et al. MUC5B Promoter Variant and Rheumatoid Arthritis with Interstitial Lung Disease. N Engl J Med. 2018. Epub 2018/10/23. doi: 10.1056/NEJMoa1801562. PubMed PMID: 30345907.

42. Seibold MA, Wise AL, Speer MC, Steele MP, Brown KK, Loyd JE, et al. A common MUC5B promoter polymorphism and pulmonary fibrosis. N Engl J Med. 2011;364(16):1503–12. Epub 2011/04/22. doi: 10.1056/NEJMoa1013660. PubMed PMID: 21506741; PubMed Central PMCID: PMCPMC3379886.

43. Armanios MY, Chen JJ, Cogan JD, Alder JK, Ingersoll RG, Markin C, et al. Telomerase mutations in families with idiopathic pulmonary fibrosis. N Engl J Med. 2007;356(13):1317–26. Epub 2007/03/30. doi: 10.1056/NEJMoa066157. PubMed PMID: 17392301.

44. Juge PA, Borie R, Kannengiesser C, Gazal S, Revy P, Wemeau-Stervinou L, et al. Shared genetic predisposition in rheumatoid arthritis-interstitial lung disease and familial pulmonary fibrosis. Eur Respir J. 2017;49(5). Epub 2017/05/13. doi: 10.1183/13993003.02314-2016. PubMed PMID: 28495692.

45. Solomon JJ, Matson S, Kelmenson LB, Chung JH, Hobbs SB, Rosas IO, et al. IgA Antibodies Directed Against Citrullinated Protein Antigens Are Elevated in Patients With Idiopathic Pulmonary Fibrosis. Chest. 2020;157(6):1513–21. Epub 2019/12/27. doi: 10.1016/j.chest.2019.12.005. PubMed PMID: 31877269; PubMed Central PMCID: PMCPMC7268435.

46. Solomon JJ, Chung JH, Cosgrove GP, Demoruelle MK, Fernandez-Perez ER, Fischer A, et al. Predictors of mortality in rheumatoid arthritis-associated interstitial lung disease. Eur Respir J. 2016;47(2):588–96. doi: 10.1183/13993003.00357-2015. PubMed PMID: 26585429.

47. Lynch DA, Godwin JD, Safrin S, Starko KM, Hormel P, Brown KK, et al. High-resolution computed tomography in idiopathic pulmonary fibrosis: diagnosis and prognosis. Am J Respir Crit Care Med. 2005;172(4):488–93. Epub 2005/05/17. doi: 10.1164/rccm.200412-1756OC. PubMed PMID: 15894598.

48. Edey AJ, Devaraj AA, Barker RP, Nicholson AG, Wells AU, Hansell DM. Fibrotic idiopathic interstitial pneumonias: HRCT findings that predict mortality. Eur Radiol. 2011;21(8):1586–93. Epub 2011/03/08. doi: 10.1007/s00330-011-2098-2. PubMed PMID: 21380740.

49. Shin KM, Lee KS, Chung MP, Han J, Bae YA, Kim TS, et al. Prognostic determinants among clinical, thin-section CT, and histopathologic findings for fibrotic idiopathic interstitial pneumonias: tertiary hospital study. Radiology. 2008;249(1):328–37. Epub 2008/08/07. doi: 10.1148/radiol.2483071378. PubMed PMID: 18682581.

50. Richeldi L, du Bois RM, Raghu G. Efficacy and safety of nintedanib in idiopathic pulmonary fibrosis. … England Journal of …. 2014. doi: 10.1056/NEJMoa1402584.

51. Kelly C, Matteson E, Aringer M, Burmester GR, Mueller H, Moros L, et al. OP0124 EFFECTS OF NINTEDANIB IN PATIENTS WITH PROGRESSIVE FIBROSING INTERSTITIAL LUNG DISEASE ASSOCIATED WITH RHEUMATOID ARTHRITIS (RA-ILD) IN THE INBUILD TRIAL. Ann Rheum Dis. 2021;80(Suppl 1):69–. doi: 10.1136/annrheumdis-2021-eular.969.

52. Behr J, Prasse A, Kreuter M, Johow J, Rabe KF, Bonella F, et al. Pirfenidone in patients with progressive fibrotic interstitial lung diseases other than idiopathic pulmonary fibrosis (RELIEF): a double-blind, randomised, placebo-controlled, phase 2b trial. Lancet Respir Med. 2021;9(5):476–86. Epub 2021/04/03. doi: 10.1016/S2213-2600(20)30554-3. PubMed PMID: 33798455.

