## Supplementary material for "A Randomized, Double-Blinded, Placebo-Controlled, Phase 2 Study of Safety, Tolerability and Efficacy of Pirfenidone in Patients with Rheumatoid Arthritis Interstitial Lung Disease": Complete supplement

Supplementary Appendix

This appendix has been supplied by the authors to give readers additional information about their work.

Contents:

Appendix A: List of Investigators

Appendix B: TRAIL1 Study Committees

Appendix C: TRAIL1 Trial Design

Appendix D: TRAIL1 Inclusion and Exclusion Criteria

Appendix E: Secondary Endpoints: OMERACT and the Dyspnea-12

Table S1: Reasons for Not Meeting Eligibility Requirements

Table S2: Subgroup Analysis of Adverse Events Among Subjects with Certain Concomitant Medications

Figure S1. Percent Predicted FVC by Patient and Treatment Group in Intention to Treat Population

Figure S2. FVC (L) by Patient and Treatment Group in Intention to Treat Population

Figures S3. Mean Percent Predicted FVC by Treatment Group in Intention to Treat Population

Figures S4. Mean FVC (L) by Treatment Group in Intention to Treat Population

Figures S5. Mean Percent Predicted FVC by Treatment Group and UIP-Pattern in Intention to Treat Population

Figures S6. Mean FVC (L) by Treatment Group and UIP-Pattern in Intention to Treat Population

**Appendix A: List of Investigators and Study Coordinators**

**(*USA*) Baylor College of Medicine, Houston, TX:** Shana Haynes-Harp, Fernando Poli, Coimbatore Sree Vidya; **Brigham and Women’s Hospital, Boston, MA:**  Rebecca R. Baron, Timothy Clouser, Tracy Doyle, Anthony Maeda; **Cleveland Clinic, Cleveland, OH:** Kristin B. Highland; **Johns Hopkins Hospital, Baltimore, MD:** Jemima F. Albayda, Sarah E. Collins, Karthik S. Suresh; **Mayo Clinic, Rochester, MN:** John M. Davis, Andrew H. Limper;  **National Jewish Health, Denver CO:**	Isabel Amigues, Kristina Eliopoulos, Jeffery J. Swigris; **National Jewish Imaging Core, Denver, CO** Stephen Humphries; **Tulane, New Orleans, LA:** John C. Huntwork, Chris Glynn, RN; **University of Alabama, Birmingham, AL:** Steve R. Duncan, Maria I. Danila; **University of Miami, Miami, FL:** Marilyn K. Glassberg, Elana M. Oberstein; **University of Michigan, Ann Arbor, MI:** Elizabeth A. Belloli, Linda Briggs, Vivek Nagaraja; **University of Michigan Data Coordination Center (DCC),** Linda Cholewa, Donna DiFranco, Edward Green, Christie Liffick, Tanvi Naik; **University of San Francisco, San Francisco, CA:** Genevieve Montas; **University of Utah, Salt Lake City, UT:** Dorota Lebiedz-Odrobina; **University of Washington, Seattle, WA:** Reba Bissell, Mark Wener; **Vanderbilt University, Vanderbilt, TN:** Lisa H. Lancaster, Leslie J. Crawford; **Weill Cornell Medicine, New York, NY:** Karmela Chan, Robert J. Kaner, Alicia Morris, Xiaoping Wu. **(*Canada*)** **McMaster University, Hamilton, ON**: Nader A. Khalidi, **University of British Columbia, Vancouver, BC:** Christopher J. Ryerson, Alyson W. Wong; **University of Calgary, Calgary, AB;** Charlene D. Fell, Sharon A. LeClercq, Mark Hyman; **University of Toronto, Toronto, ON:** Shane Shapera, Shikha Mittoo; **(*United Kingdom*)** **Glenfield Hospital, Leicester, UK:** Shireen Shaffu, **Norfolk and Norwich University Hospital, Norwich, UK:** Karl Gaffney; Andrew M. Wilson; **North Bristol NHS Trust:** Shaney Barratt, Harsha Gunawardena; **Oxford University Hospitals, Oxford, UK:** Rachel K. Hoyles, Joel David, Namrata Kewalramani; **Royal Brompton Hospital, London, UK:** Toby M. Maher, Philip L. Molyneaux, Maria A. Kokosi; **Royal Devon and Exeter Hospital, Exeter, UK:** Matthew J. Cates, Jessica Mandizha; **Royal Edward Infirmary, Wigan, UK:** Abdul Ashish, Gladstone Chelliah; **Royal Papworth Hospital, Cambridgeshire, UK:** Helen Parfrey, Muhunthan Thillai; **Royal Victoria Infirmary, Newcastle upon Tyne, UK:** Ian Forrest, Josephine Vila; **Southampton General Hospital, Southampton, UK:** Sophie V. Fletcher; **St James’s University Hospital, Leeds, UK:** Paul Beirne; Clair Favager; **University Hospital Aintree, Liverpool, UK:** Jo Brown, Julie K. Dawson; **Wythenshawe Hospital, Manchester, UK**: Pilar Rivera Ortega, Sahena Haque, Pippa Watson. **(*Australia*) Alfred Health, Melbourne:** Jun K. Khoo, Karen Symons; **Royal Prince Alfred Hospital, Camperdown:** Peter Youssef; **The Prince Charles Hospital, The University of Queensland, Chermside:** John A. Mackintosh.

**Appendix B - TRAIL1 Study Committees**

**Clinical Coordinating Committee (CCC)**

**Brigham and Women’s Hospital/Baylor College of Medicine**
Ivan Rosas, MD (Study PI)

Hilary Goldberg, MD (CCC Director)

Shana Haynes-Harp, BS, MSA, DHSc (Project Manager, US)

Shelley Hurwitz, PhD (Lead Statistician)

Rie Mauer, MS (Statistician)

Donna Walsh (Administrative Operations)

Timothy Clouser, MBA (Finance Manager)

Donna DiFranco (QA Director)

**United Kingdom (UK) Sponsor**

Felix Woodhead, MD, PhD (UK PI)

Sarah Terry (Project Manager, UK)

Victoria Harris (Project Manager, UK)

**Canadian Sponsor**

**Australian Sponsor**

Daniel C. Chambers, MD (Australian PI)

**Data Coordinating Center (DCC)**

**University of Michigan**

Jen Mawby, RN, CCRP (Project Manager)

Cathie Spino, DSc (DCC Director)

Linda Cholewa, CIP (Clinical Monitor)

Tanvi Naik, MS (Project Manager)

**Clinical & Regulatory Leads and Advisor**

Sonye Danoff, MD (Regulatory Lead)

Joshua J. Solomon, MD (Clinical Lead)

Mark A. Perrella, MD (Medical Monitor)

**Imaging Core**

David Lynch, MB, BCh

Stephen Humphries, PhD

Martin Kolb, MD, PhD (Canadian PI)

**Appendix C: TRAIL1 Trial Design**

Visit 1 (week -8 to week 0) SCREENING

- Obtain informed consent
- Screen potential subjects by inclusion and exclusion criteria
- H&P, labs, ECG, HRCT, Spirometry, DLCO, biomarkers (see study assessment calendar for details)

Visit 2 (week 0) ELIGIBILITY/RANDOMIZATION

- H&P, labs, PROs, joint count, Spirometry, biomarkers (see study assessment calendar for details)
- Confirm eligibility and randomize patient
- Dispense study drug
- AE assessment

Visit 3 (week 1) TELEPHONE ASSESSMENT

- Vital status, dosing compliance
- Concomitant medication assessment
- AE assessment

Visit 4 (week 2) TELEPHONE ASSESSMENT

- Vital status, dosing compliance
- Concomitant medication assessment
- AE assessment

Visit 5 (week 4) and Visit 6 (week 8) PROTOCOL VISIT

- H&P, labs, biomarkers (week 4 only), ECG (week 4 only)
- Review drug compliance
- AE assessment

Visit 7 (week 13), Visit 9 (week 26) and Visit 10 (week 39) PROTOCOL VISIT / DISPENSE DRUG

- H&P, labs, biomarkers, PROs, ECG, Spirometry
- Dispense supply of study drug
- AE assessment

Visit 8 (week 19) LAB ASSESSMENT

- Liver function testing
- Concomitant medication assessment
- AE assessment

Visit 11 (week 52) END OF TREATMENT

- H&P, labs, biomarkers, PROs, joint count, Spirometry, DLCO, ECG, HRCT
- Collect diary and unused study drug
- AE assessment

Visit 12 END OF STUDY PHONE CALL (28 DAYS AFTER LAST DOSE OF STUDY DRUG)

- Vital status and AE assessment

PRE-RESTART VISIT

- H&P, labs, ECG
- AE assessment
- Dispense supply of study drug, if required
- Review drug compliance, dispense new diary, if required

UNSCHEDULED VISIT

- H&P, AE assessment
- remaining elements of the visit are up to the discretion of the investigator

EARLY TERMINATION VISIT

- H&P, labs, biomarkers, PROs, joint count , Spirometry, DLCO, ECG, HRCT (see study assessment calendar for details)
- Collect diary and unused study drug
- AE assessment

**Appendix D: TRAIL1 Inclusion and Exclusion Criteria**

**Inclusion**

1. Age 18 through 85 years, inclusive, at Screening
2. Probable or definite diagnosis of RA according to revised 2010 ACR/EULAR criteria, without evidence or suspicion of an alternative diagnosis that may contribute to their interstitial lung disease.
3. Diagnosis of ILD
   a. supported by clinically indicated HRCT, and when available, surgical lung biopsy (SLB), prior to Screening, and
   b. presence of fibrotic abnormality affecting more than 10% of the lung parenchyma, with or without traction bronchiectasis or honeycombing, on Screening and confirmed by adjudicated HRCT prior to Baseline
4. No features supporting an alternative diagnosis on transbronchial biopsy, or SLB, if performed prior to Screening
5. Attainment of the following centralized spirometry criteria (based on local spirometry on standardized equipment and centralized quality controlled):
   a. percent predicted FVC ≥ 40% at Screening
   b. change in pre-bronchodilator FVC (measured in liters) between Screening (Visit 1) and Baseline (Visit 2) must be a <10% relative difference, calculated as: 100% * [absolute value (Screening FVC – Baseline FVC) / Screening FVC ]
   c. percent predicted DLCO or TLCO ≥30 % at Screening
   d. Screening (Visit 1) pre-bronchodilator(BD) and Post-BD spirometry meets ATS quality criteria as determined by a central reviewer
   e. Baseline (Visit 2) Pre-BD spirometry meets ATS quality criteria as determined by the site Investigator or the central reviewer
6. Able to understand and sign a written informed consent form
7. For women of childbearing potential: agreement to remain abstinent (refrain from heterosexual intercourse) or use two adequate methods of contraception, including at least one method with a failure rate of <1% per year, during the 52-week treatment period and for at least 118 days after the last dose of study drug
   a. A woman is considered to be of childbearing potential if she is postmenarcheal, has not reached a postmenopausal state (≥ 12 continuous months of amenorrhea with no identified cause other than menopause), and has not undergone surgical sterilization (removal of ovaries and/or uterus).
   b. Examples of contraceptive methods with a failure rate of <1% per year include bilateral tubal ligation, male sterilization, established and proper use of hormonal contraceptives that inhibit ovulation, hormone-releasing intrauterine devices, and copper intrauterine devices.
   c. The reliability of sexual abstinence should be evaluated in relation to the duration of the clinical trial and the preferred and usual lifestyle of the patient. Periodic abstinence (e.g., calendar, ovulation, symptothermal, or postovulation methods) and withdrawal are not acceptable methods of contraception.
8. For men who are not surgically sterile: agreement to remain abstinent (refrain from heterosexual intercourse) or use contraceptive measures, and agreement to refrain from donating sperm, as defined below:
   a. With female partners of childbearing potential, men must remain abstinent or use a condom plus an additional contraceptive method that together result in a failure rate of < 1% per year during the treatment period and for at least 118 days after the last dose of study drug.
   b. Men must refrain from donating sperm during this same period.

**Exclusion**

1. Not a suitable candidate for enrollment or unlikely to comply with the requirements of this study, in the opinion of the investigator
2. Cigarette smoking within 3 months of Screening or unwilling to avoid tobacco products throughout the study
3. History of clinically significant environmental exposure known to cause pulmonary fibrosis (PF), including but not limited to drugs (such as amiodarone), asbestos, beryllium, radiation, and domestic birds
4. Concurrent presence of the following conditions:
   a. Other interstitial lung disease, related to but not limited to radiation, drug toxicity, sarcoidosis, hypersensitivity pneumonitis, or bronchiolitis obliterans organizing pneumonia
   b. Medical history including Human Immunodeficiency Virus (HIV)
   c. Medical history of viral hepatitis (positive Hep A antibody in the absence of elevated liver enzymes is not an exclusion)
5. Concurrent presence of other pleuropulmonary manifestations of RA, including but not limited to rheumatoid nodular disease of the lung, pleuritis/pleural thickening, and obliterative bronchiolitis
6. Post-bronchodilator FEV1/FVC < 0.7 at Screening
7. Presence of pleural effusion occupying more than 20% of the hemithorax on Screening HRCT
8. Clinical diagnosis of a second connective tissue disease or overlap syndrome (including but not limited to scleroderma, Sjogren’s, polymyositis/dermatomyositis, systemic lupus erythematosus but excluding Raynaud’s phenomena)
9. Coexistent clinically significant COPD/emphysema or asthma in the opinion of the site principal investigator
10. Clinical evidence of active infection, including but not limited to bronchitis, pneumonia, sinusitis, urinary tract infection, or cellulitis. The infection should be resolved per PI assessment prior to enrollment. Any use of antibiotics must be completed 2 weeks prior to the screening visit. Note that prophylactic antibiotics are not contraindicated or exclusionary
11. Any history of malignancy diagnosed within 5 years of screening, other than basal cell carcinoma of the skin, squamous cell carcinoma of the skin, or low-grade cervical carcinoma in situ
12. History of LFT abnormalities as outlined below, or imaging, laboratory or other clinical information suggesting liver dysfunction, advanced liver disease or cirrhosis.  Evidence of hepatic impairment that in the opinion of the investigator could interfere with drug metabolism or increase the risk of the known hepatotoxicity of study drug. Any of the following liver function abnormalities:
    a. Total bilirubin above the upper limit of normal (ULN), excluding patients with Gilbert’s syndrome
    b. Aspartate or alanine aminotransferase (AST/SGOT or AST/SGPT) > 3 X ULN
    c. Alkaline phosphatase > 2.5 X ULN
13. History of end-stage renal disease requiring dialysis
15. History of unstable or deteriorating cardiac disease, or unstable cardiac arrhythmia or arrhythmia requiring modification of drug therapy, myocardial infarction within the previous year, heart failure requiring hospitalization. Any condition that, in the opinion of the investigator, might be significantly exacerbated by the known side effects associated with the administration of pirfenidone
16. History of alcohol or substance abuse in the past 2 years, at the time of Screening
17. Family or personal history of long QT syndrome
18. Any of the following test criteria above specified limits:
    a. Estimated glomerular filtration rate <30 mL/min/1.73m2
    b. ECG with a QTc interval >500 msec at Screening
19. Prior use of pirfenidone or known hypersensitivity to any of the components of study treatment
20. Use of any of the following therapies within 28 days before Screening and during participation in the study:
    a. Investigational therapy, defined as any drug that has not been approved for marketing for any indication in the country of the participating site
    b. Potent inhibitors of CYP1A2 (e.g., fluvoxamine, enoxacin)
    c. Potent inducers of CYP1A2
    d. Sildenafil (daily use). Note: intermittent use for erectile dysfunction is allowed
21. Introduction and/or modification of dose of corticosteroids or any cytotoxic, immunosuppressive, or cytokine modulating or receptor antagonist agent for the management of **pulmonary** manifestations of RA, within 3 months of screening, **is** an exclusion criterion for enrollment, with the exception of dose modification of systemic corticosteroids that are maintained at or below 20 mg prednisone daily or the equivalent. However, introduction and/or modification of dose of corticosteroids or any cytotoxic, immunosuppressive, or cytokine modulating or receptor antagonist agent for the management of **extrapulmonary** manifestations of RA **is not** an exclusion criterion for enrollment.
22. Any use of an approved anti-fibrotic medication within 28 days of screening.

**Appendix E: Secondary Endpoints: OMERACT and the Dyspnea-12**

An international interdisciplinary network of experts in ILD was formed to propose and select outcomes in randomized clinical trials in ILD. A working group of this network looking at outcome measures in CTD-ILD (the Outcome Measures in Rheumatology (OMERACT) initiative) defined “clinically meaningful progression” in CTD-ILD as a relative decline from baseline in percent predicted FVC of ≥10%, or relative decline from baseline in percent predicted FVC ≥ 5% and <10%, and ≥15% relative decline in DLCO_._ The Dyspnea-12 is comprised of 12 breathless descriptors that are scored on a 4-item scale. The item scores are summed and can be divided into a Physical and Affective domain. It has shown good internal consistency and repeatability in patients with ILD and is simple to complete.

**Table S1: Reasons for not meeting eligibility requirements**

| **Reasons** | **N (%)** |
| --- | --- |
| INC2. RA diagnosis | 2 (1.9) |
| INC3A. ILD diagnosis via HRCT and/or SLB | 7 (6.5) |
| INC3B. ILD diagnosis 6 months prior to screening | 3 (2.8) |
| INC3C. ILD diagnosis fibrotic abnormality on HRCT | 2 (1.9) |
| INC5A. Spirometry percent predicted FVC greater than or equal to 40% and less than or equal to 100% at Screening | 14 (13.1) |
| INC5B. Spirometry change in pre-bronchodilator FVC (measured in liters) between V1 and V2 must be a < 10% relative difference | 11 (10.3) |
| INC5C. Spirometry percent predicted DLCO greater than or equal to 30% and less than or equal to 100% at Screening | 12 (11.2) |
| INC6A. Spirometry percent predicted FVC greater than or equal to 40% and less than or equal to 80% at Screening | 7 (6.5) |
| INC6C. Spirometry percent predicted DLCO greater than or equal to 30% and less than or equal to 80% at Screening | 7 (6.5) |
| INC7. Inclusion criteria: Stable dose RA meds | 1 (0.9) |
| EXC1. Not suitable candidate for study | 13 (12.2) |
| EXC2. Cigarette smoking | 1 (0.9) |
| EXC3. Environmental exposure | 1 (0.9) |
| EXC4C. Presence of viral hepatitis | 1 (0.9) |
| EXC5. Presence of other pleuropulmonary RA | 1 (0.9) |
| EXC6. Post-bronchodilator FEV1/FVC <0.7 at Screening | 9 (8.4) |
| EXC8. Clinical diagnosis of a second connective tissue disease or overlap syndrome | 1 (0.9) |
| EXC10. Clinical evidence of active infection | 8 (7.5) |
| EXC11. Any history of malignancy diagnosed within 5 years of screening | 2 (1.9) |
| EXC12. History of LFT abnormalities or evidence of hepatic impairment | 1 (0.9) |
| EXC14. History of unstable or deteriorating cardiac or disease | 4 (3.7) |
| EXC18B. ECG out of range | 1 (0.9) |
| EXC19A. Bilirubin out of range | 1 (0.9) |

**Table S2: Subgroup Analysis of Adverse Events Among Subjects with Concomitant Medications (anti-inflammatories, TNF agents, and rituximab)**

|  | Pirfenidone (N=56) | Placebo (N=48) | P value |
| --- | --- | --- | --- |
| Treatment-emergent adverse events  Treatment-emergent serious adverse events | 56 (100.0%)  9 (16.1%) | 46 (95.8%)  6 (12.5%) | 0.2106  0.6053 |

**Figure S1. Percent Predicted FVC by Patient and Treatment Group in Intention to Treat Population**

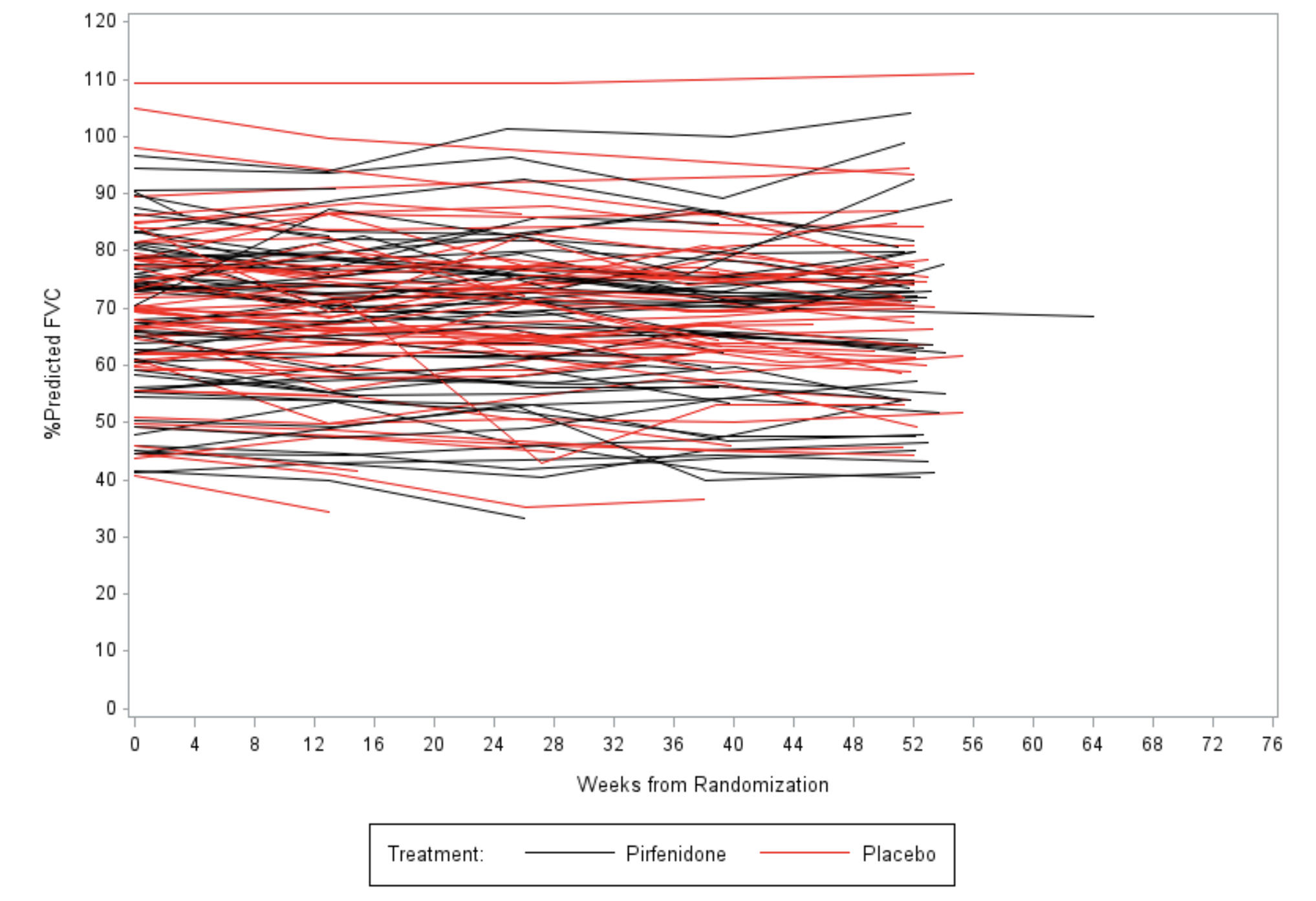

**Figure S2. FVC (L) by Patient and Treatment Group in Intention to Treat Population**

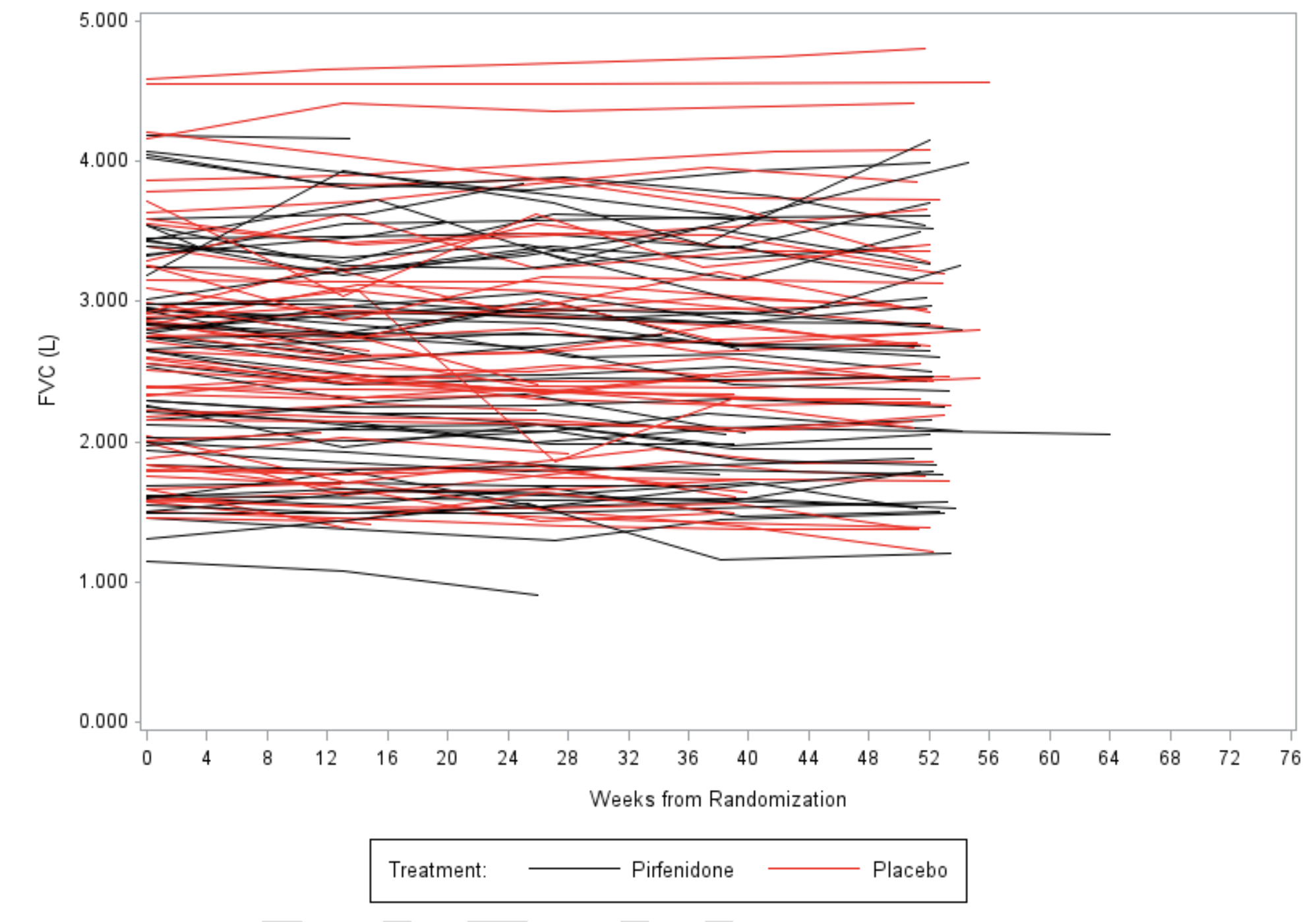

**Figures S3. Mean Percent Predicted FVC by Treatment Group in Intention to Treat Population**

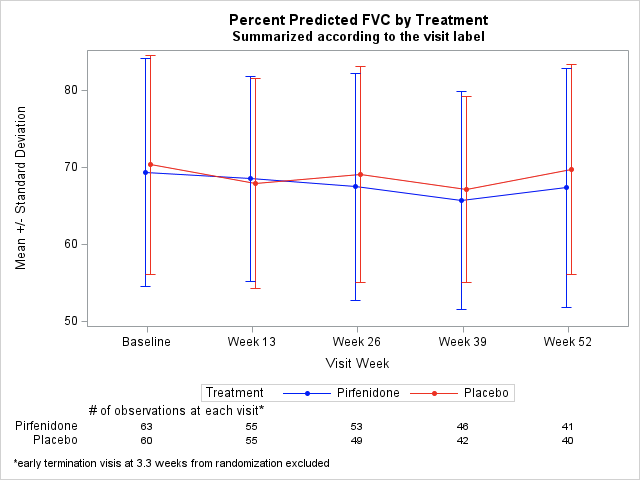

**Figures S4. Mean FVC (L) by Treatment Group in Intention to Treat Population**

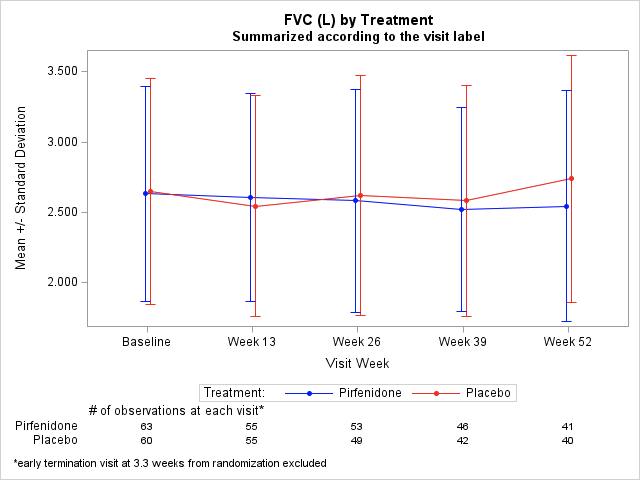

**Figures S5. Mean Percent Predicted FVC by Treatment Group and UIP-Pattern in Intention to Treat Population**

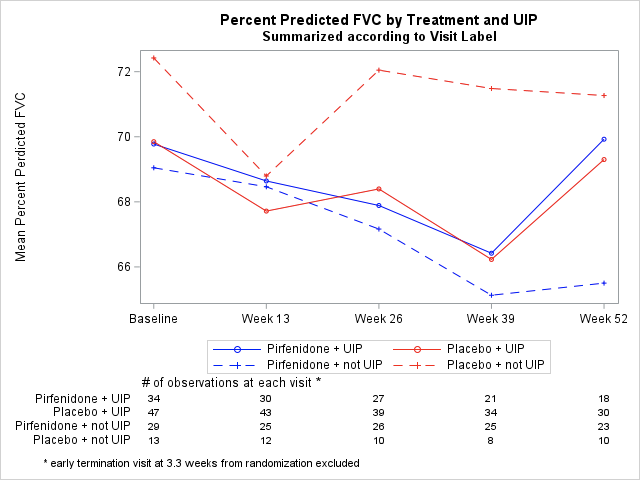

**Figures S6. Mean FVC (L) by Treatment Group and UIP-Pattern in Intention to Treat Population**

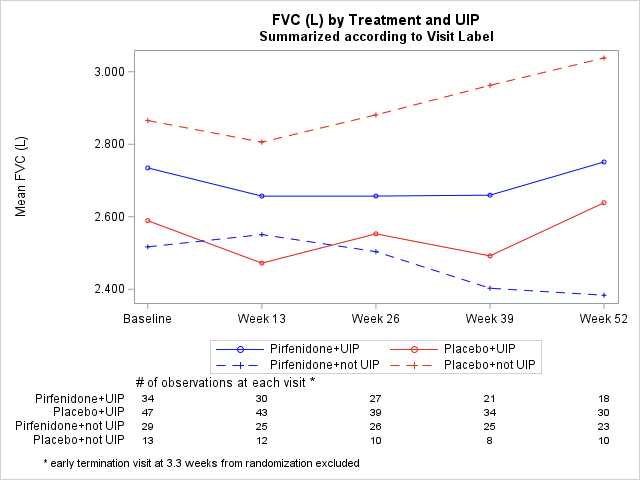
